# Metformin mitigates insulin signaling variations induced by COVID-19 vaccine boosters in type 2 diabetes

**DOI:** 10.1101/2023.12.27.23299358

**Authors:** Lixiang Zhai, Min Zhuang, Hoi Ki Wong, Chengyuan Lin, Jialing Zhang, Gengyu Bao, Yijing Zhang, Shujun Xu, Jingyuan Luo, Shuofeng Yuan, Hoi Leong Xavier Wong, Zhao-xiang Bian

## Abstract

Diabetes is associated with an increased risk of Coronavirus disease 2019 (COVID-19) vulnerability and mortality. COVID-19 vaccines significantly reduce the risks of serious COVID-19 outcomes, but the impact of COVID-19 vaccines including their effectiveness and adverse effects in patients with diabetes are not well known yet. Here, we showed that 61.1% patients with type 2 diabetes, but not healthy controls, exhibited aggravated insulin resistance towards the booster shots of the COVID-19 vaccine. Furthermore, we showed that COVID-19 vaccination once a week also impaired insulin sensitivity in healthy mice after four weeks. We further showed that metformin, a common anti-diabetic medication, improved the impaired insulin signaling induced by COVID-19 vaccination in mice. This study suggests clinical implications for the close monitoring of glycemic control in diabetic patients after receiving COVID-19 vaccines and indicates the beneficial action of metformin in counteracting insulin signaling variations induced by COVID-19 vaccination in diabetic patients.

## Introduction

Diabetes mellitus (DM), commonly referred to as diabetes, is a chronic metabolic disorder characterized by high levels of glucose in the blood, either due to insufficient insulin production (type 1 diabetes) or insulin resistance (type 2 diabetes). With approximately 10% of the global population affected by this condition, diabetes has become one of the most prevalent metabolic diseases worldwide[1]. The impact of diabetes extends beyond the direct implications of the disease itself, as it predisposes individuals to a myriad of complications[2]. Over time, diabetes can lead to a cascade of health complications. Cardiovascular diseases, for instance, are notably more common among those with diabetes, as the condition contributes to the development of atherosclerosis, increasing the risk of heart attacks and strokes. Diabetic neuropathy is another frequent complication, where high blood sugar levels cause damage to the nerve fibers, particularly in the legs and feet, leading to numbness, tingling, or pain. Nephropathy, which is kidney damage resulting from diabetes, can progress to kidney failure, a life-threatening condition.

Coronavirus disease 2019 (COVID-19), a global pandemic caused by severe acute respiratory syndrome coronavirus 2 (SARS-CoV-2), has brought challenges for people with diabetes as they have an increased risk for more severe symptoms and complications from COVID-19[3]. Diabetic patients are more susceptible to an array of complications from the SARS-CoV-2, including a higher likelihood of requiring intensive care and mechanical ventilation, and a greater risk of death compared to the general population[4]. As diabetes is associated with an impaired immune response, infection with COVID-19 can lead to more severe and prolonged illness in diabetic patients.

COVID-19 vaccination has been shown to reduce the risk of symptomatic infection, hospitalization rates, and mortality from COVID-19[5]. Moreover, large-scale vaccination data have demonstrated a substantial decrease in hospitalization rates and mortality due to COVID-19 among the diabetic population[6]. The collective data strongly suggest that the benefits of receiving a COVID-19 vaccine significantly outweigh the risks, particularly for individuals with pre-existing health conditions such as diabetes. Herein COVID-19 vaccination has become a critical defense for individuals with diabetes. By bolstering the immune response to SARS-CoV-2, vaccines help prevent the hyper-inflammatory reactions and severe respiratory complications that can arise from a COVID-19 infection.

Recognizing the advantages of the COVID-19 vaccine, it is also imperative to elucidate any unforeseen effects, especially in populations with pre-existing metabolic conditions. Meanwhile, as many countries have used booster doses including third and fourth doses of COVID-19 vaccine to maintain immune protection[7], there is a need for ongoing surveillance to track any adverse outcomes of COVID-19 vaccination in individuals with diabetes. As the interplay between COVID-19, vaccination, and diabetes is not entirely understood, it is critical to conduct both clinical and basic scientific research to understand COVID-19 vaccine impact on diabetes.

## Results

### COVID-19 vaccine aggravates insulin resistance in diabetic patients

We first conducted a longitudinal clinical trial to determine the effects of the COVID-19 vaccine in healthy controls, pre-diabetic subjects, and diabetic subjects. Between 1 June 2023 and 31 October 2023, we recruited 155 adults who have received two doses of the mRNA COVID-19 vaccine (BNT162b2). The participants ranged in age from 18 to 65 years (median=53.5 years, IQR 12.0) and 60.25% are male. Human volunteers were recruited to determine their immune responses and glucose control before and 2 weeks after the booster shot of COVID-19 mRNA vaccination. As expected, we showed SARS-CoV-2 spike protein, IgG antibodies against SARS-CoV-2 spike (Trimer) and neutralizing abilities of SARS-CoV-2 protein significantly increased after treatment of COVID-19 mRNA booster in both healthy controls and diabetic patients as shown by levels of SARS-CoV-2 spike protein, measurement of IgG antibodies against SARS-CoV-2 spike (Trimer) and SARS-CoV-2 surrogate virus neutralization test (sVNT) (p<0.001 in all cases, Figure.1A-C), but no significant differences are found in immune responses indexes among healthy controls, pre-diabetic subjects and diabetic subjects (n.s. in all cases, Figure.1A-C), suggesting diabetic patients exhibited similar immune responses as healthy controls towards COVID-19 mRNA booster.

**Figure1.**
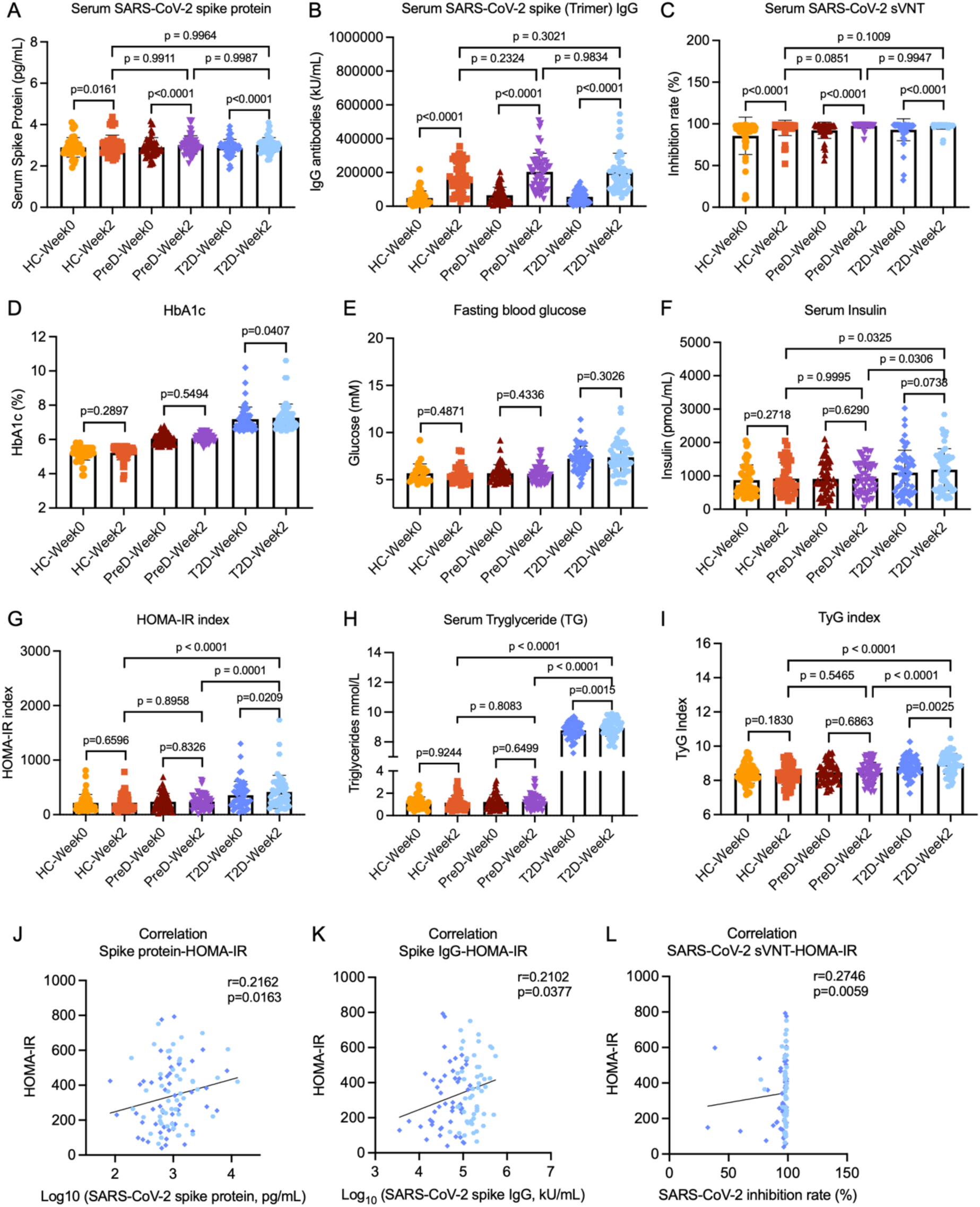
COVID-19 vaccine aggravates insulin resistance in diabetic subjects. **(A-C)** SARS-CoV-2 spike protein, SARS-CoV-2 spike (Trimer) IgG and SARS-CoV-2 sVNT in serum samples from healthy controls (HC), pre-diabetic subjects (PreD) and diabetic subjects (T2D) following vaccination of BNT mRNA COVID-19 vaccine (n=56 per group) (determined by two-tailed t-tests). **(D-I)** HbA1c, fasting blood glucose (FBG), fasting insulin, homeostatic model assessment for insulin resistance (HOMA-IR), triglyceride (TG) and triglyceride-glucose (TyG) index in serum samples from diabetic subjects following vaccination of BNT mRNA COVID-19 vaccine (n=56 per group) (determined by two-tailed t-tests). **(J-L)** Spearman’s correlation between serum SARS-CoV-2 spike protein, SARS-CoV-2 spike (Trimer) IgG and SARS-CoV-2 sVNT index with HOMA-IR index in diabetic subjects following vaccination of BNT mRNA COVID-19 vaccine (n=56 per group) (determined by two-tailed t-tests).

However, in contrast to immune responses to COVID-19 mRNA boosters shown in healthy controls, pre-diabetic subjects and diabetic subjects, we found exacerbated risks of glucose intolerance and insulin resistance after the booster shots of COVID-19 mRNA vaccination in pre-diabetic patients and diabetic patients, as revealed by the significant elevation of HbA1c, homeostatic model assessment for insulin resistance (HOMA-IR), triglyceride (TG) and triglyceride-glucose index (TyG) (p<0.02 in all cases, Figure.1D and G-I). In contrast, no significant changes in fasting blood glucose (FBG) and insulin levels are found in human subjects after the booster shots of COVID-19 mRNA vaccination (n.s. in all cases, Figure.1E-F). About 61.1% of diabetic subjects had impairment of insulin sensitivity according to the HOMA-IR index and about 66.7% of diabetic subjects had increased risks of cardiovascular complications according to the TyG index. Moreover, correlation analysis revealed HOMA-IR index is positively correlated with a series of immune responses including SARS-CoV-2 spike protein, SARS-CoV-2 spike (Trimer) IgG, and neutralizing abilities of SARS-CoV-2 protein after COVID-19 vaccination (p<0.05 in all cases, r=0.2162, 0.2102 and 0.2746, Figure.1J-L). These results suggest that the booster shot of mRNA COVID-19 vaccine impairs glucose control and aggravates insulin resistance in human subjects with type 2 diabetes.

### COVID-19 vaccine impairs insulin signaling in mice

To study the inhibitory action of COVID-19 vaccination on glucose control, we first performed a glucose tolerance test in healthy mice that received mRNA COVID-19 vaccine (BNT162b2) weekly. Compared to mice treated with saline, mice treated with the COVID-19 vaccine exhibited immune responses similar as shown by the elevation of SARS-CoV-2 spike protein in serum (p=0.001, Figure.2A). Interestingly, we showed mice after the fourth dose of COVID-19 vaccine exhibited impaired glucose tolerance examined by oral glucose tolerance test (OGTT) (p<0.05 in all cases, Figure.2B-C). We also showed serum triglyceride, but not FBG, serum insulin level or bodyweight, is significantly elevated in mice with weekly COVID-19 vaccination (p=0.0305 for Figure.2D, n.s. for Figure.2E-G), indicating an increased risk of cardiovascular diseases and metabolic disorders. Coupled with impaired glucose tolerance, the reduction in blood glucose in response to insulin challenge in the insulin tolerance test (ITT) (p<0.05 in all case, Figure.2H-I) and the level of insulin-induced Akt phosphorylation in insulin-sensitive tissues were significantly reduced in mice with COVID-19 vaccination (p<0.05 in all cases, Figure.2J-M). These results suggested that the glucose intolerance induced by the COVID-19 vaccine is mediated by impairment of insulin sensitivity rather than impaired insulin secretion in mice.

**Figure.2.**
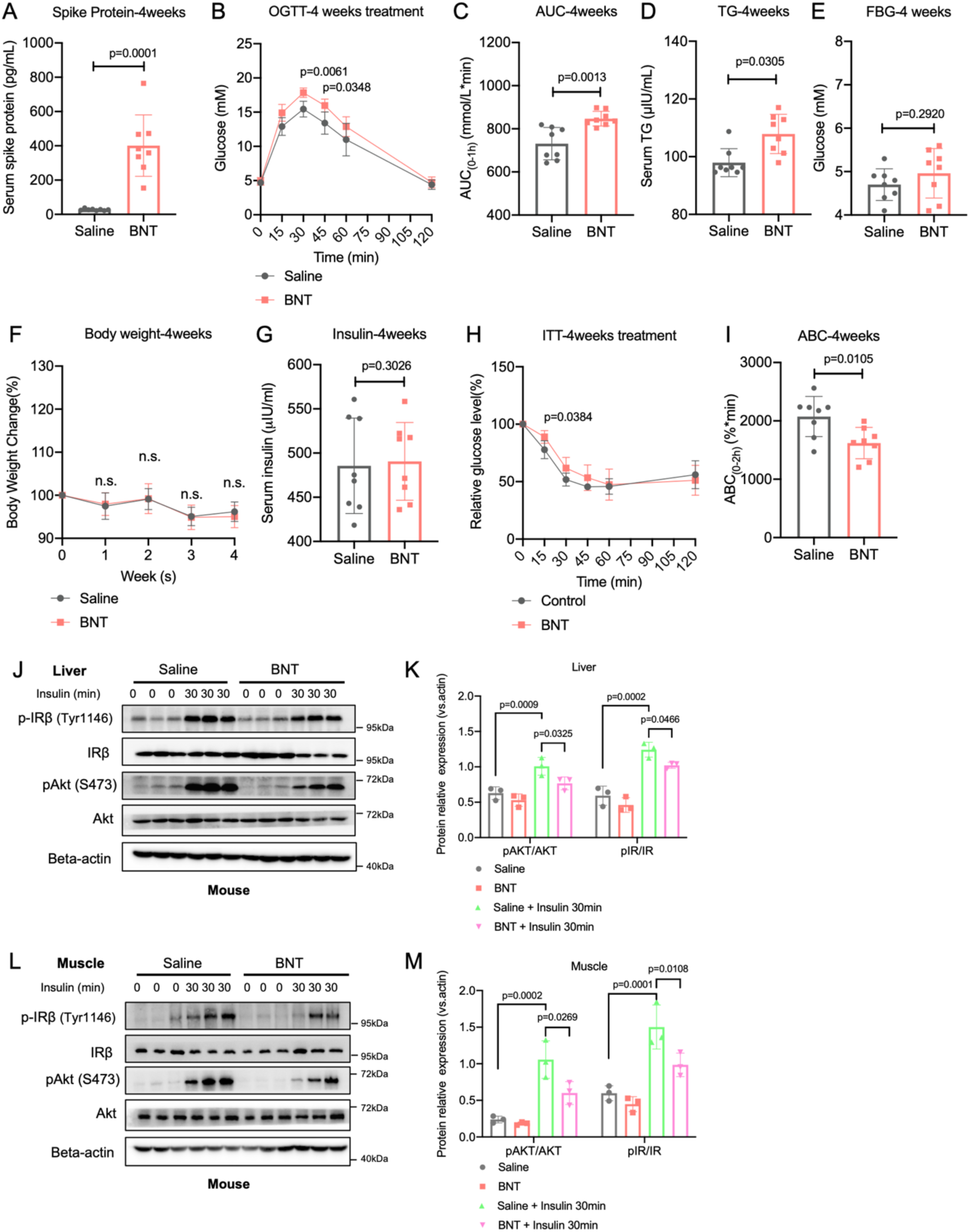
COVID-19 vaccine impairs insulin sensitivity in mice. **(A)** SARS-CoV-2 spike protein level, fasting TG (triglyceride) and FBG (fasting blood glucose) levels in serum samples from normal mice following treatment of BNT mRNA COVID-19 vaccine (4.5 μg/kg) for 4 weeks (n=8 per group) (determined by two-tailed t-tests). **(B-C)** OGTT and AUC (area under curve) indexes in normal mice following treatment of BNT mRNA COVID-19 vaccine (4.5 μg/kg) for 4 weeks (n=8 per group) (determined by two-tailed t-tests). **(D-G)** Fasting TG (triglyceride), FBG (fasting blood glucose), fasting insulin levels in serum samples and bodyweight index from normal mice following treatment of BNT mRNA COVID-19 vaccine (4.5 μg/kg) for 4 weeks (n=8 per group) (determined by two-tailed t-tests). **(H-I)** ITT and ABC (area below curve) indexes in normal mice following treatment of BNT mRNA COVID-19 vaccine for 4 weeks (4.5 μg/kg) (n=8 per group) (determined by two-tailed t-tests). **(J-M)** Western blot (and quantification) of the effects of BNT mRNA COVID-19 vaccine (4.5 μg/kg) on IR (insulin receptor) and AKT phosphorylation stimulated by insulin (1U/kg) in liver and muscle lysates from normal mice (n=3/group) (determined by two-tailed one-way ANOVA test).

### Metformin alleviates insulin resistance induced by COVID-19 vaccine in mice

Metformin, a first-line anti-diabetes drug, has been found to improve the COVID-19 severity and mortality[13], and reduce the incidence of post-acute COVID-19 syndrome (PACS)[14], whereas the beneficial effects of metformin on the COVID-19 induced insulin resistance remain elusive. Interestingly, we demonstrated that metformin at a clinically relevant dosage alleviated insulin resistance in mice vaccinated with the COVID-19 vaccine as shown by OGTT and ITT indexes (p<0.05 in all cases, Figure.3A-D) and rescued the impaired insulin signaling in mice (p<0.05 in all cases, Figure.3E-H). Notably, metformin did not affect the protective effects of COVID-19 vaccine against SARS-CoV-2 as demonstrated by the non-significant changes in serum levels of SARS-CoV-2 spike protein, spike protein IgG and SARS-CoV-2 sVNT (n.s. in all cases, Figure.4I-K). These results suggest metformin can be used as an adjunctive treatment for maintaining glucose control after COVID-19 vaccination.

**Figure.3.**
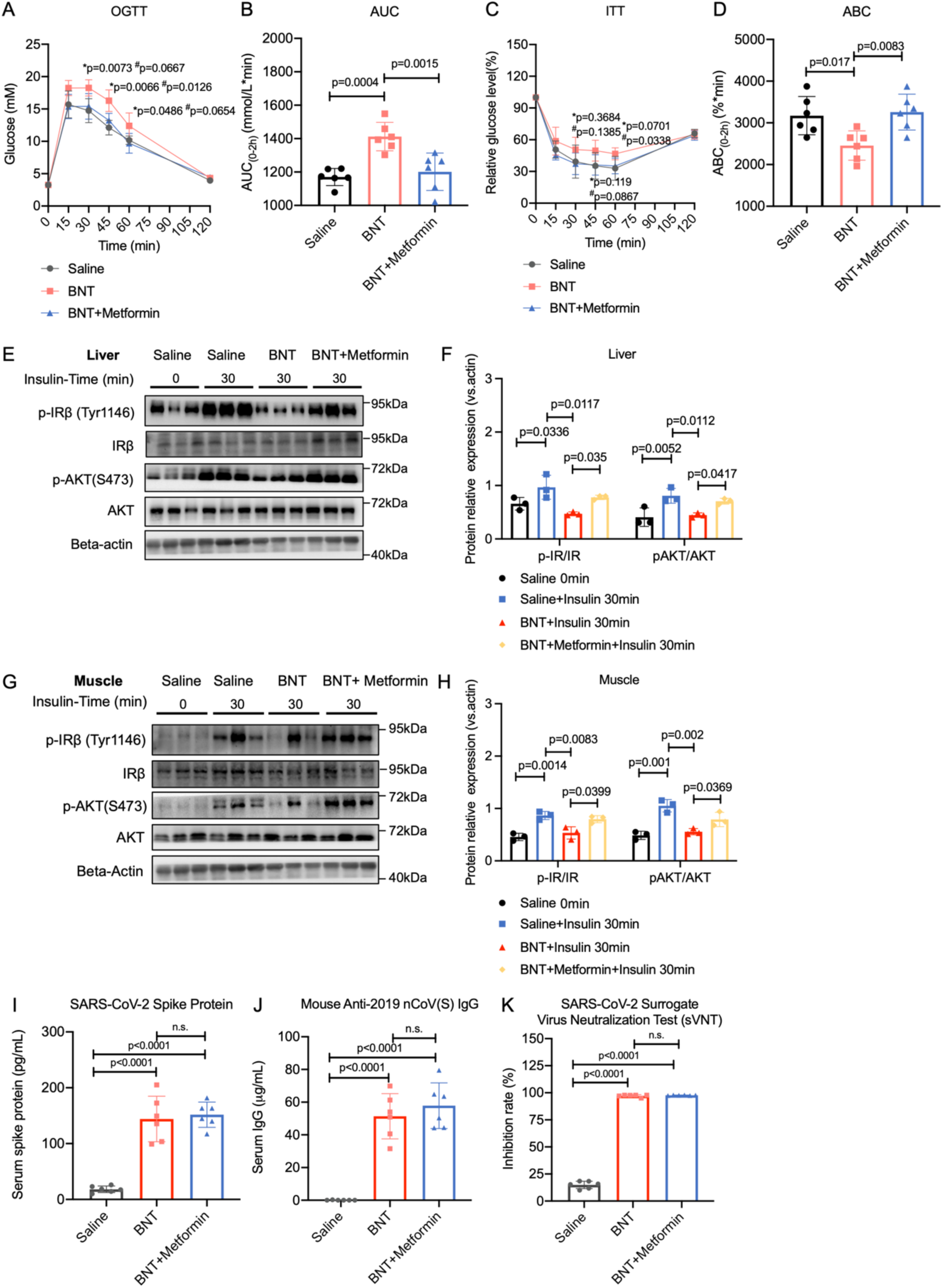
Metformin alleviates COVID-19 vaccine-induced insulin resistance. **(A-B)** OGTT and AUC indexes in normal mice following treatment of BNT mRNA COVID-19 vaccine (4.5 μg/kg) and metformin (300mg/kg) (n=6 per group) (determined by two-tailed t-tests). **(C-D)** ITT and ABC indexes in normal mice following treatment of BNT mRNA COVID-19 vaccine (4.5 μg/kg) and metformin (300mg/kg) (n=6 per group) (determined by two-tailed t-tests). **(E-H)** Western blot (and quantification) of the effect of BNT mRNA COVID-19 vaccine (4.5 μg/kg) and metformin (300mg/kg) on IR and AKT phosphorylation (n=6/group) in liver and muscle lysates (determined by two-tailed one-way ANOVA test). **(I-K)** SARS-CoV-2 spike protein, SARS-CoV-2 spike (Trimer) IgG and SARS-CoV-2 sVNT in in serum samples from normal mice following treatment of BNT mRNA COVID-19 vaccine (4.5 μg/kg) and metformin (300mg/kg) (n=3 per group) (determined by two-tailed one-way ANOVA test).

## Discussion

The rapid development and widespread distribution of COVID-19 vaccines have played a crucial role in mitigating the global impact of the pandemic. While these vaccines have proven to be highly effective in preventing severe COVID-19 illness and reducing transmission rates, the long-term effects of COVID-19 vaccines have not been well characterized yet[7]. In this study, we showed that boosters of COVID-19 vaccines weaken insulin sensitivity in pre-diabetic and diabetic patients, providing a causal link between COVID-19 vaccines and insulin resistance. Several studies have been conducted to assess the impact of COVID-19 vaccines on glucose metabolism and insulin sensitivity[4, 16]. Although these studies found no significant changes in glucose tolerance as measured by fasting blood glucose following single-time vaccination, we showed that the booster shots of the COVID-19 vaccine impair insulin sensitivity and increase diabetic complications risks in diabetic patients as measured by risk factors of insulin resistance and biological indexes of insulin sensitivity. Moreover, we showed multiple times (>3) COVID-19 vaccinations significantly impair insulin signaling in healthy mice accompanied by elevation of SARS-CoV-2 spike protein and IgG antibodies of SARS-CoV-2 spike protein, resulting in impaired glucose control and insulin signaling. As studies showed post-acute COVID-19 syndrome (PACS) linked to the persistence of SARS-CoV-2 spike protein and IgG antibodies of SARS-CoV-2 spike protein, patients with PACS may also have impaired insulin signaling and increased risks of diabetes and its complications.

Recent studies have uncovered the association between COVID-19 and the risk of T2D. A large retrospective cohort study found that patients with a history of COVID-19 had a significantly higher risk of developing T2D within one year of infection compared to the general population[17]. Additionally, a systematic review and meta-analysis reported an increased risk of new-onset diabetes in individuals who had recovered from COVID-19[18]. In addition, we revealed a common anti-diabetes medication metformin, which is associated with improved clinical outcomes of COVID-19 and reduced risks of developing PACS-related symptoms by clinical studies[14], significantly improved insulin sensitivity induced by COVID-19 vaccines in mice without affecting the levels of SARS-CoV-2 spike protein. Our results also delivered basic and clinical implications of metformin for the treatment of COVID-19 and PACS-related insulin resistance in subjects with pre-diabetes and diabetes.

There are several potential mechanisms through which boosters of COVID-19 might increase the risk of T2D development in diabetic patients. First, SARS-CoV-2 spike protein induced by booster of COVID-19 vaccine may contribute to insulin resistance and impaired glucose metabolism[3]. The induction of SARS-CoV-2 spike protein may persist even after several months of the COVID-19 vaccination, affecting the development of T2D in susceptible individuals. SARS-CoV-2 spike protein induced by boosters of the COVID-19 vaccine may directly affect insulin signaling via binding toll-like receptor 4 (TLR4) and estrogen receptor (ER) which is involved in the regulation of insulin signaling [19], which may lead to reduced insulin secretion and insulin resistance. Second, the SARS-CoV-2 spike protein induced by the COVID-19 vaccine is known to cause systemic immune responses and the production of IgG antibodies of the SARS-CoV-2 spike protein by the host, which may affect the function of insulin signaling pathways and lead to insulin resistance. We also noticed that the insulin sensitivity and triglyceride levels were only affected by boosters of COVID-19 in pre-diabetic and diabetic subjects but not healthy controls in our present study, suggesting subjects with impaired glucose tolerance are recommended to take extra care of their blood glucose homeostasis after COVID-19 vaccination. A long-term longitudinal study in the future will help better monitor the glucose control and complications risks in pre-diabetic and diabetic patients with boosters of the COVID-19 vaccine.

## Data Availability

All data produced in the present study are available upon reasonable request to the authors

## Acknowledgments

This study was kindly supported by Health@InnoHK Initiative Fund from the Hong Kong SAR Government (ITC RC/IHK/4/7 to ZX.B.), Key-Area Research and Development Program of Guangdong Province (2020B1111110003 to ZX.B.), NSFC Excellent Young Scientist Scheme (32322091 to HLX.W.), Innovation and Technology Support Programme from the Hong Kong SAR Government (ITS/058/22MS, to HLX.W.) and General Research Grants from the Hong Kong SAR Government (22104123, to HLX.W.). We also thank all volunteers who participated in this study.

## Author contributions

Conceptualization: ZX.B., HLX.W. and LX.Z. Methodology: HLX.W., LX.Z., M.Z. and CY.L. Investigation: M.Z., GY.B., YJ.Z., SJ.X. HK.W., JL.Z., JY.L. Visualization: M.Z. and LX.Z. Funding acquisition: ZX.B. Project administration: ZX.B., HLX.W., LX.Z. and CY.L. Supervision: ZX.B., HLX.W. and LX.Z. Writing-original draft: M.Z. and LX.Z. Writing-review & editing: ZX.B., HLX.W., LX.Z and SF.Y.

## Declaration of interests

The authors declare that they have no competing interests.

## Materials and methods

The experiments were not randomized, and investigators were not blinded to allocation during experiments and outcome assessment.

### Reagents and resources

Reagents and resource details are provided in Table.S1.

### Human subjects

The human study was approved by the Research Committee on the Use of Human and Animal Subjects in Teaching and Research at Hong Kong Baptist University (REC/22-23/0277) and registered on Chinese Clinical Trial Registry (ChiCTR2300069830). A total of 180 participants receiving BioNTech (BNT) mRNA COVID-19 vaccination (60 in each of the following groups: pre-diabetes mellitus, diabetes mellitus and healthy controls) are recruited and observed over a 2-week period for their changes in insulin sensitivity before and after the vaccination of BNT to determine the effects of spike protein on insulin sensitivity.

### Animals

The mice study was approved by the Research Committee on the Use of Human and Animal Subjects in Teaching and Research at Hong Kong Baptist University (REC/22-23/0611). The mice experiments were performed under the regulation of the animal ordinance of the Department of Health, Hong Kong SAR, China. Male BALB/c mice (6-8 weeks and 20-25g), which were purchased from the Laboratory Animal Services Center, Chinese University of Hong Kong, were raised in the Animal Unit, School of Chinese Medicine, Hong Kong Baptist University. The mice were kept at a condition of 12/12 h light/dark cycle at a controlled temperature of approximately 25°C with free access to food and water.

Mice were administered either with BioNTech COVID-19 mRNA vaccine by intramuscular injection at a dosage of 4.5 μg/kg according to the adjustment of dosage used in human once per week for 5 weeks. Mice were administered with related agonists or antagonists of TLR4, ER and ACE2, respectively in the mechanism study.

### Oral glucose tolerance test (OGTT) and insulin glucose tolerance test (ITT)

For OGTT, mice were fasted overnight (12 hours) before the OGTT and given glucose solution at a dosage of 2g/kg orally and blood samples were collected from the caudal vein for glucose measurement at 0, 15, 30, 45, 60, 90 and 120 min after the glucose administration. The area under the concentration-time curve (AUC) was calculated. For ITT, mice were fasted for 4 hours before the ITT and given insulin by i.p. at a dosage of 1U/kg and blood samples were collected from the caudal vein for glucose measurement at 0, 15, 30, 45, 60, 90 and 120 min as per OGTT. The area above the concentration-time curve (ABC) was also calculated.

### Serological tests

Mice were fasted overnight (12 hours) before the collection of serum samples. Serum samples (about 200 μL) were collected by orbital bleeding in mice under anesthesia conditions. SARS-CoV-2 spike RBD IgG test, SARS-CoV-2 surrogate virus neutralization test (sVNT), SARS-CoV-2 spike protein ELISA test, triglyceride and insulin levels were measured according to the protocols provided by the manufacturer.

### Western blotting

Frozen mice tissue samples and harvested cells were lysed in RIPA buffer and normalized to a concentration of 2mg/mL protein. For western blotting, tissue lysates and cell lysates were mixed with 5x loading buffer and heated at 98 ℃ for 10 min. The target proteins were detected as per the instruction protocols of BioRad. The blots were then incubated with HRP-linked anti-rabbit IgG or anti-mouse IgG and reacted with chemiluminescence. The semi-quantification of blots was analyzed with Image J.

## Statistical analysis

Data was expressed as average and SD values in at least triplicates. *P*-value was calculated using Graphpad Prism 9 and *p*-values less than 0.05 are considered statistically significant. Unpaired student’s t-tests or one-way ANOVA analysis were employed in analysis settings as indicated.

## Data and materials availability

Further information and requests for clinical resources and information can be directed to Lixiang Zhai. This study does not generate new experimental materials.

## Notes

### Competing Interest Statement

The authors have declared no competing interest.

### Clinical Trial

ChiCTR2300069830

### Author Declarations

The human study was approved by the Research Committee on the Use of Human and Animal Subjects in Teaching and Research at Hong Kong Baptist University (REC/22-23/0277) and registered on Chinese Clinical Trial Registry (ChiCTR2300069830).

